# Symptoms and symptom clusters associated with SARS-CoV-2 infection in community-based populations: Results from a statewide epidemiological study

**DOI:** 10.1101/2020.10.11.20210922

**Authors:** Brian E. Dixon, Kara Wools-Kaloustian, William F. Fadel, Thomas J. Duszynski, Constantin Yiannoutsos, Paul K. Halverson, Nir Menachemi

## Abstract

**Background:** Prior studies examining symptoms of COVID-19 are primarily descriptive and measured among hospitalized individuals. Understanding symptoms of SARS-CoV-2 infection in pre-clinical, community-based populations may improve clinical screening, particularly during flu season. We sought to identify key symptoms and symptom combinations in a community-based population using robust methods.

**Methods:** We pooled community-based cohorts of individuals aged 12 and older screened for SARS-CoV-2 infection in April and June 2020 for a statewide seroprevalence study. Main outcome was SARS-CoV-2 positivity. We calculated sensitivity, specificity, positive predictive value (PPV), and negative predictive value (NPV) for individual symptoms as well as symptom combinations. We further employed multivariable logistic regression and exploratory factor analysis (EFA) to examine symptoms and combinations associated with SARS-CoV-2 infection.

**Results:** Among 8214 individuals screened, 368 individuals (4.5%) were RT-PCR positive for SARS-CoV-2. Although two-thirds of symptoms were highly specific (>90.0%), most symptoms individually possessed a PPV <50.0%. The individual symptoms most greatly associated with SARS-CoV-2 positivity were fever (OR=5.34, p<0.001), anosmia (OR=4.08, p<0.001), ageusia (OR=2.38, p=0.006), and cough (OR=2.86, p<0.001). Results from EFA identified two primary symptom clusters most associated with SARS-CoV-2 infection: (1) ageusia, anosmia, and fever; and (2) shortness of breath, cough, and chest pain. Moreover, being non-white (13.6% vs. 2.3%, p<0.001), Hispanic (27.9% vs. 2.5%, p<0.001), or living in an Urban area (5.4% vs. 3.8%, p<0.001) was associated with infection.

**Conclusions:** Symptoms can help distinguish SARS-CoV-2 infection from other respiratory viruses, especially in community or urgent care settings where rapid testing may be limited. Symptoms should further be structured in clinical documentation to support identification of new cases and mitigation of disease spread by public health. These symptoms, derived from asymptomatic as well as mildly infected individuals, can also inform vaccine and therapeutic clinical trials.

**Research in Context:** *Evidence before this study:* Using multiple journal articles queried from MEDLINE as well as a Cochrane systematic review, we examined all studies that described symptoms known to be associated with COVID-19. We further examined the guidelines from WHO and CDC on the symptoms those public health authorities consider to be associated with COVID-19. Most of the evidence comes from China, Italy, and the United States. Collectively prior research and guidance suggests there are a dozen symptoms reported by individuals who tested positive for COVID-19 in multiple countries. Symptoms include fever, cough, fatigue, anosmia, ageusia, shortness of breath, chills, myalgias, headache, sore throat, chest pain, and gastrointestinal issues. The evidence is generally of low quality as it is descriptive in nature, and it is biased towards hospitalized patients. Most studies report the proportion of patients hospitalized or testing positive for infection who report one or more symptoms within 3-14 days prior to hospitalization or infection. There has been little validation of symptoms among hospitalized or non-hospitalized patients. Furthermore, according to a Cochrane review, no studies to date assess combinations of different signs and symptoms.

*Added value of this study:* This study employs multiple, rigorous methods to examine the ability of specific symptoms as well as symptom combinations/groups to predict laboratory-confirmed (RT-PCR) infection of SARS-CoV-2. Furthermore, the study is unique in its large sample drawn exclusively from community-based populations rather than hospitalized patients.

*Implication of all the available evidence:* Combining the evidence from this study with prior research suggests that anosmia and ageusia are key symptoms that differentiate COVID-19 from influenza-like symptoms. Clinical screening protocols for COVID-19 should look for these symptoms, which are not commonly asked of patients who present to urgent care or hospital with flu-like symptoms.

**Key points:** Important symptoms specific to COVID-19 are fever, anosmia, ageusia, and cough. Two-thirds of symptoms were highly specific (>90.0%), yet most symptoms individually possessed a PPV <50.0%. This study confirms using robust methods the key symptoms associated with COVID-19 infection, and it also identifies combinations of symptoms strongly associated with positive infection

## INTRODUCTION

Severe acute respiratory syndrome coronavirus 2 (SARS-CoV-2) causes COVID-19 disease, which has a range of manifestations from asymptomatic infection to severe pneumonia, potentially leading to intensive care utilization or death. Globally there have been more than 23 million cases and over 808 000 deaths as of August 24, 2020. Despite the advent of summer when incidence of most respiratory viruses decreases, COVID cases increased in the U.S. Given that the upcoming fall-winter season in the northern hemisphere typically increases influenza circulation, it is important to understand the common and differentiating symptoms of these two diseases.

Symptoms of COVID-19 are similar to those of other influenza-like illnesses. Based primarily on studies of hospitalized individuals, the U.S. Centers for Disease Control and Prevention (CDC) recognized three principal symptoms for COVID-19: fever, cough, and shortness of breath.^1^ This list was expanded as the pandemic progressed to include chills, myalgias, headache, sore throat, and the loss of taste (ageusia) and/or smell (anosmia). An early systematic review that included 1,576 hospitalized COVID-19 patients reported that the most prevalent clinical symptom was fever, followed by cough, fatigue and dyspnea.^2^ A later review reported the main clinical symptoms to be fever, cough, fatigue, slight dyspnea, sore throat, headache, conjunctivitis and gastrointestinal issues.^3^ In a community-based study involving self-reported symptoms via a mobile app, 10 symptoms -- fever, persistent cough, fatigue, shortness of breath, diarrhea, delirium, skipped meals, abdominal pain, chest pain and hoarse voice – were associated with self-reported positive test results in a UK cohort.^4^ A Cochrane systematic review ^5^ identified a total of 27 signs and symptoms for COVID-19.

Importantly, much of the information about prevalent COVID-19 symptoms originate from studies focused on limited populations who presented in a hospital setting.^5^ Moreover, testing guidelines in the US, and many other parts of the world, have prioritized symptomatic and high-risk individuals, which further biases available data on symptomology towards those with more severe disease. Most studies further focus exclusively on SARS-CoV-2 positive patients and thus lack an uninfected control group which might also exhibit some level of baseline symptoms. In Menni et al.,^4^ app users self-reported test results limiting reliability of findings in an uncontrolled study. The Cochrane review suggests the existing evidence on symptoms is “highly variable,” and no studies to date assessed combinations of different signs and symptoms.^5^ In summary, there is much that can still be learned about the symptomatology of SARS-CoV-2 infection, especially among community-based individuals who may not require or have not yet presented for clinical care.

We sought to examine the symptoms reported by populations of community-based individuals tested for SARS-CoV-2 in the context of a statewide prevalence study. We examine patterns and groups of symptoms, and we compare individuals who tested positive for active viral infection, using RT-PCR, to those who screened negative. We also examine symptom differences by age, sex, race, ethnicity, and rurality. By better characterizing COVID-19 symptoms, especially among those that may have milder disease, we sought to better identify those symptoms and symptom combinations that are more likely to represent SARS-CoV-2 infection. In addition, understanding the symptomology of community-based infections has the potential to inform the selection of end points for consideration in clinical trials focused on vaccine and therapeutic effectiveness.

## METHODS

### Study Participants and Recruitment

Data derived from two waves of testing in Indiana, conducted in partnership with the state health department, were analyzed for this study. Each wave included groups that were either randomly selected or that sought testing in open-testing sites throughout the state. Wave 1 of testing, described elsewhere,^6^ occurred at the end of April 2020. Wave 2 occurred in the beginning of June 2020. The random sample for each wave was selected from individual state tax records of filers and dependents. In addition, because underrepresented minority groups are more seriously impacted by COVID-19, both waves also conducted targeted nonrandom testing, in conjunction with religious and civic leaders, in select African American and Hispanic communities. In all cases, inclusion criteria were Indiana residency and being 12 years of age or older. Individuals from both random and nonrandom samples were tested regardless of symptoms, prior testing history, or medical history. Testing was available with no out-of-pocket costs to participants.

Recruitment was aided by public announcements by the Governor, the media, and minority community leaders. State agencies, including the Indiana Minority Health Coalition, reached out to minority communities to encourage participation. The testing was a component of the state’s overall effort to expand testing capacity and surveillance for COVID-19.

### COVID-19 Testing and Specimen Collection

Using Dacron swabs and standard techniques, trained personnel collected nasopharyngeal swabs for RT-PCR analysis. Nasopharyngeal swabs were transferred to the laboratories of Eli Lilly and Company (Lilly Clinical Diagnostics Lab SARS-CoV-2 test based on the CDC primary set) or Indiana University Health (Luminexon NxTAG CoV Extended Panel or Roche cobas SARS-CoV-2 test) for RT-PCR testing. All laboratories were located in Indianapolis, IN. Test results were reported to participants via a secure website within 1 to 4 days.

### Data Collection

Upon arrival to a testing site, each participant was asked to complete a research intake form that included questions about symptoms, health status, and demographics. Using a standardized checklist, participants were asked to indicate presence of symptoms within the past two weeks (14 days). The checklist represented a composite of symptoms reported by either the WHO or the CDC to be associated with COVID-19^1^ and expanded to include additional symptoms reported in the literature.^7,8^ Participants were requested to identify all symptoms present, and those who did not indicate any symptoms were categorized as asymptomatic.

### Statistical Analysis

Because the target populations, identification, and recruitment processes were similar between the two waves, all participants from both waves were pooled for analysis. Descriptive statistics were calculated for all individuals who participated in testing for SARS-CoV-2, stratified by the recruitment method (e.g., random, or nonrandom). Chi-square tests were used to compare various characteristics between the two groups as well as characteristics of individuals testing positive for SARS-CoV-2 versus those testing negative.

Using all available data, we first examined the sensitivity, specificity, positive predictive value (PPV), and negative predictive value (NPV) of each symptom reported by participants. Because specific combinations of symptoms might be useful in screening patients who present to a clinic or hospital, we further explored various symptom combinations. First, we examined the sensitivity, specificity, PPV and NPV of symptom pairs and triplets. All such symptom permutations were examined. To further explore symptom combinations, we employed exploratory factor analysis (EFA) to create symptom groups using positive RT-PCR status as the gold standard for identification of SARS-CoV-2 infection. Factor analysis is a recommended multivariate method for exploring symptoms when symptoms are commonly grouped together in a given etiology.^9^

Furthermore, we developed a logistic regression model to examine the relationship between the presence of individual symptoms and positive RT-PCR status, controlling for participant demographics and sample recruitment method. The logistic regression model computes the probability of positive RT-PCR status ranging from 0 to 1. We then generated a Receiver Operating Characteristic (ROC) curve associated with the model by varying the cut-off probability across the range of observed values using the pROC package in R. These are plots of the true-positive (sensitivity) versus the false-positive rate (1 – specificity) of a test, over all possible cutoff points.^10^ We also calculated the area under the ROC curve (AUC) as a global measure of the accuracy of the model to predict SARS CoV-2 positivity.

All analyses were performed using R (version 3.6.3). This study was determined to be exempt by the Institutional Review Board (IRB) at Indiana University under the public health surveillance exception.

### Role of the Funding Source

The funder of the study had no role in study design, data collection, data analysis, data interpretation, or writing of the paper. The corresponding author had full access to all the data in the study and had final responsibility for the decision to submit for publication.

## RESULTS

We screened a total of 8214 individuals for SARS-CoV-2 including 6326 (77.0%) individuals who were randomly selected and 1888 (23.0%) who were recruited for nonrandom testing in partnership with racial and ethnic minority community leaders. A total of 368 individuals (4.5%) were RT-PCR positive for SARS-CoV-2. The characteristics of tested participants are summarized in **Table 1**. Randomly selected and nonrandom participants were significantly (p<0.001) different on all characteristics. The nonrandom group included more males (45% vs. 41%, p<0.001), non-whites (57.2% vs. 8.5%, p<0.001), Hispanics (27% vs. 2.3%, p<0.001), and urban (80.6% vs. 65%, p<0.001) residents. Males and females tested positive in similar proportions (4.6% vs. 4.4%, p=0.707). Non-whites (13.6% vs. 2.3%, p<0.001), Hispanics (27.9% vs. 2.5%, p<0.001), and individuals living in Urban areas (5.4% vs. 3.8%, p<0.001) had higher rates of positivity.

**Table 1.** Characteristics of Indiana residents who were tested for SARS-CoV-2 infection in late April and early June 2020, includes are individuals randomly selected for testing along with nonrandom samples designed to enhance inclusion of minority communities.

| **Characteristics** | **Total**<br>**Participants**<br>**No. (Col %)** | **Randomly**<br>**Selected**<br>**Individuals**<br>**No. (Col %)** | **Individuals**<br>**from Open**<br>**Community**<br>**Testing**<br>**Locations**<br>**No. (Col %)** | **P Value<sup>a</sup>**<br>**for**<br>**Group** | **Individuals**<br>**Testing**<br>**Positive via**<br>**RT-PCR**<br>**No. (Row**<br>**%)** | **P Value<sup>b</sup>**<br>**for**<br>**Positivity** |
| --- | --- | --- | --- | --- | --- | --- |
| Overall Population Totals | 8214 | 6326 | 1888 |  | 368 |  |
| Female | 4565 (55.6%) | 3451 (54.6%) | 1114 (59.0%) | <0.001 | 201 (4.4%) | 0.71 |
| Male | 3647 (44.4%) | 2873 (45.4%) | 774 (41.0%) |  | 167 (4.6%) |  |
| White | 6599 (80.3%) | 5791 (91.5%) | 808 (42.8%) | <0.001 | 149 (2.3%) | <0.001 |
| Non-white | 1615 (19.7%) | 535 (8.5%) | 1080 (57.2%) |  | 219 (13.6%) |  |
| Hispanic | 653 (7.9%) | 144 (2.3%) | 509 (27.0%) | <0.001 | 182 (27.9%) | <0.001 |
| Non-Hispanic | 7561 (92.1%) | 6182 (97.7%) | 1379 (73.0%) |  | 186 (2.5%) |  |
| Urban<sup>c</sup> | 5631 (68.6%) | 4109 (65.0%) | 1522 (80.6%) | <0.001 | 303 (5.4%) | <0.001 |
| Rural/Mixed | 1598 (19.5%) | 1500 (23.7%) | 98 (5.2%) |  | 27 (1.7%) |  |
| Rural | 980 (11.9%) | 717 (11.3%) | 263 (13.9%) |  | 37 (3.8%) |  |
| Age: <40 | 2379 (29.0%) | 1747 (27.6%) | 632 (33.5%) | <0.001 | 167 (7.0%) | <0.001 |
| Age: 40-59 | 3036 (37.0%) | 2308 (36.5%) | 728 (38.6%) |  | 155 (5.1%) |  |
| Age: 60+ | 2799 (34.1%) | 2271 (35.9%) | 528 (28.0%) |  | 46 (1.6%) |  |

<sup>a</sup>Comparison of Randomly Selected individuals to nonrandom community testing
<sup>b</sup>Comparison of Individuals Testing Positive to those Testing Negative
<sup>c</sup>Based upon Purdue Rural Indiana Classification System

Symptoms experienced by individuals who tested positive or negative and the sensitivity, specificity, PPV, and NPV for each individual symptom with respect to predicting RT-PCR positivity are summarized in **Table 2**. Also presented are combinations of symptoms with a PPV ≥60.0%. Although two-thirds of the symptoms were highly specific for COVID-19 (>90.0%), most symptoms individually possessed a PPV of <50.0%. The three symptoms with the largest individual PPVs were anosmia (52.5%), ageusia (51.0%), and fever (47.6%). When fever was paired with anosmia or ageusia, with or without the presence of a third symptom, the PPV increased to >70%. However, sensitivity for all individual symptoms and symptom combinations was low (<50.0%).

**Table 2.** Self reported symptoms by participants undergoing SARS-CoV-2 testing in a statewide prevalence study. All individual symptoms and lack of symptoms (asymptomatic) included as well as combinations of symptoms with a positive predictive value >60%.

| Symptoms reported in the past 14 days | Number of individuals testing positive via RT-PCR<br>N (%) | Number of individuals testing negative via RT-PCR<br>N (%) | Sensitivity | Specificity | Positive Predictive Value | Negative Predictive Value |
| --- | --- | --- | --- | --- | --- | --- |
| Overall Population Totals | 368 (4.6) | 7650 (95.4) |  |  |  |  |
| Asymptomatic (no reported symptoms) | 91 (24.7) | 4681 (61.2) | 24.7% | 38.8% | 1.9% | 91.5% |
| **Individual Symptoms** |  |  |  |  |  |  |
| Loss of Smell (anosmia) | 94 (25.5) | 85 (1.1) | 25.5% | 98.9% | 52.5% | 96.5% |
| Loss of Taste (ageusia) | 105 (28.5) | 101 (1.3) | 28.5% | 98.7% | 51.0% | 96.6% |
| Fever | 129 (35.1) | 142 (1.9) | 35.1% | 98.1% | 47.6% | 96.9% |
| Chills | 81 (22) | 197 (2.6) | 22.0% | 97.4% | 29.1% | 96.3% |
| Chest Pain | 65 (17.7) | 255 (3.4) | 17.7% | 96.6% | 20.3% | 96.0% |
| Vomiting | 12 (3.3) | 51 (0.7) | 3.3% | 99.3% | 19.0% | 95.5% |
| Muscle Ache (myalgia) | 117 (31.8) | 564 (7.4) | 31.8% | 92.6% | 17.2% | 96.6% |
| Cough | 175 (47.6) | 969 (12.7) | 47.6% | 87.3% | 15.3% | 97.2% |
| Shortness of Breath | 64 (17.4) | 438 (5.8) | 17.4% | 94.2% | 12.7% | 95.9% |

| Symptoms reported in the past 14 days | Number of individuals testing positive via RT-PCR N (%) | Number of individuals testing negative via RT-PCR N (%) | Sensitivity | Specificity | Positive Predictive Value | Negative Predictive Value |
| --- | --- | --- | --- | --- | --- | --- |
| Sore Throat | 90 (24.5) | 618 (8.1) | 24.5% | 91.9% | 12.7% | 96.2% |
| Diarrhea | 59 (16) | 479 (6.3) | 16.0% | 93.7% | 11.0% | 95.8% |
| Fatigue | 127 (34.5) | 1063 (14) | 34.5% | 86.0% | 10.7% | 96.4% |
| Headache | 144 (39.1) | 1376 (18.1) | 39.1% | 81.9% | 9.5% | 96.5% |
| Runny Nose | 90 (24.5) | 1116 (14.7) | 24.5% | 85.3% | 7.5% | 95.9% |
| **Symptom Combinations** |  |  |  |  |  |  |
| Fever & Loss of Taste (ageusia) | 63 (17.1) | 24 (0.3) | 17.1% | 99.7% | 72.4% | 96.1% |
| Fever & Loss of Smell (anosmia) | 53 (14.4) | 22 (0.3) | 14.4% | 99.7% | 70.7% | 96.0% |
| Loss of Taste & Vomiting | 8 (2.2) | 4 (0.1) | 2.2% | 99.9% | 66.7% | 95.5% |
| Cough & Fever & Loss of Taste (ageusia) | 49 (13.3) | 13 (0.2) | 13.3% | 99.8% | 79.0% | 96.0% |
| Cough & Fever & Loss of Smell (anosmia) | 41 (11.1) | 13 (0.2) | 11.1% | 99.8% | 75.9% | 95.9% |
| Fever & Loss of Smell | 37 (10.1) | 13 (0.2) | 10.1% | 99.8% | 74.0% | 95.8% |

| **Symptoms reported in the past 14 days** | **Number of individuals testing positive via RT-PCR N (%)** | **Number of individuals testing negative via RT-PCR N (%)** | **Sensitivity** | **Specificity** | **Positive Predictive Value** | **Negative Predictive Value** |
| --- | --- | --- | --- | --- | --- | --- |
| (anosmia) & Muscle Ache (myalgia) |  |  |  |  |  |  |
| Fever & Loss of Taste (ageusia) & Muscle Ache (myalgia) | 46 (12.5) | 18 (0.2) | 12.5% | 99.8% | 71.9% | 95.9% |
| Fever & Loss of Smell (anosmia) & Loss of Taste (ageusia) | 45 (12.2) | 18 (0.2) | 12.2% | 99.8% | 71.4% | 95.9% |
| Fever & Headache & Loss of Smell (anosmia) | 42 (11.4) | 17 (0.2) | 11.4% | 99.8% | 71.2% | 95.9% |
| Fatigue & Fever & Loss of Smell (anosmia) | 36 (9.8) | 15 (0.2) | 9.8% | 99.8% | 70.6% | 95.8% |
| Chills & Fever & Loss of Smell (anosmia) | 26 (7.1) | 11 (0.1) | 7.1% | 99.9% | 70.3% | 95.7% |
| Diarrhea & Fever & Loss of Taste (ageusia) | 23 (6.2) | 10 (0.1) | 6.2% | 99.9% | 69.7% | 95.7% |
| Chills & Fever & Loss of | 36 (9.8) | 16 (0.2) | 9.8% | 99.8% | 69.2% | 95.8% |

| **Symptoms reported in the past 14 days** | **Number of individuals testing positive via RT-PCR**<br>**N (%)** | **Number of individuals testing negative via RT-PCR**<br>**N (%)** | **Sensitivity** | **Specificity** | **Positive Predictive Value** | **Negative Predictive Value** |
| --- | --- | --- | --- | --- | --- | --- |
| Taste (ageusia) |  |  |  |  |  |  |
| Fever & Headache & Loss of Taste (ageusia) | 47 (12.8) | 21 (0.3) | 12.8% | 99.7% | 69.1% | 95.9% |
| Fatigue & Fever & Loss of Taste (ageusia) | 42 (11.4) | 19 (0.2) | 11.4% | 99.8% | 68.9% | 95.9% |

### Symptom Groups

The principal symptom groups identified through EFA are summarized in **Figure 1**. Five principal symptom groups emerged, all of which have face validity. The first symptom group (Factor 1) consists of ageusia (r=0.92), anosmia (r=0.90), fever (r=0.63). The second symptom group (Factor 2) consists of shortness of breath (r=0.83), cough (r=0.49), and chest pain (r=0.64). The third symptom group (Factor 3) consists of fatigue (r=0.73) and myalgias (r=0.71). The fourth symptom group (Factor 4) consists of vomiting (r=0.90) and diarrhea (r=0.55). The final symptom group (Factor 5) consists of runny nose (r=0.80) and sore throat (r=0.44). The cumulative variance explained by all symptom groups was 70%, with the first two groups (Factors 1 and 2) explaining 49% of the variance.

**Figure 1.**
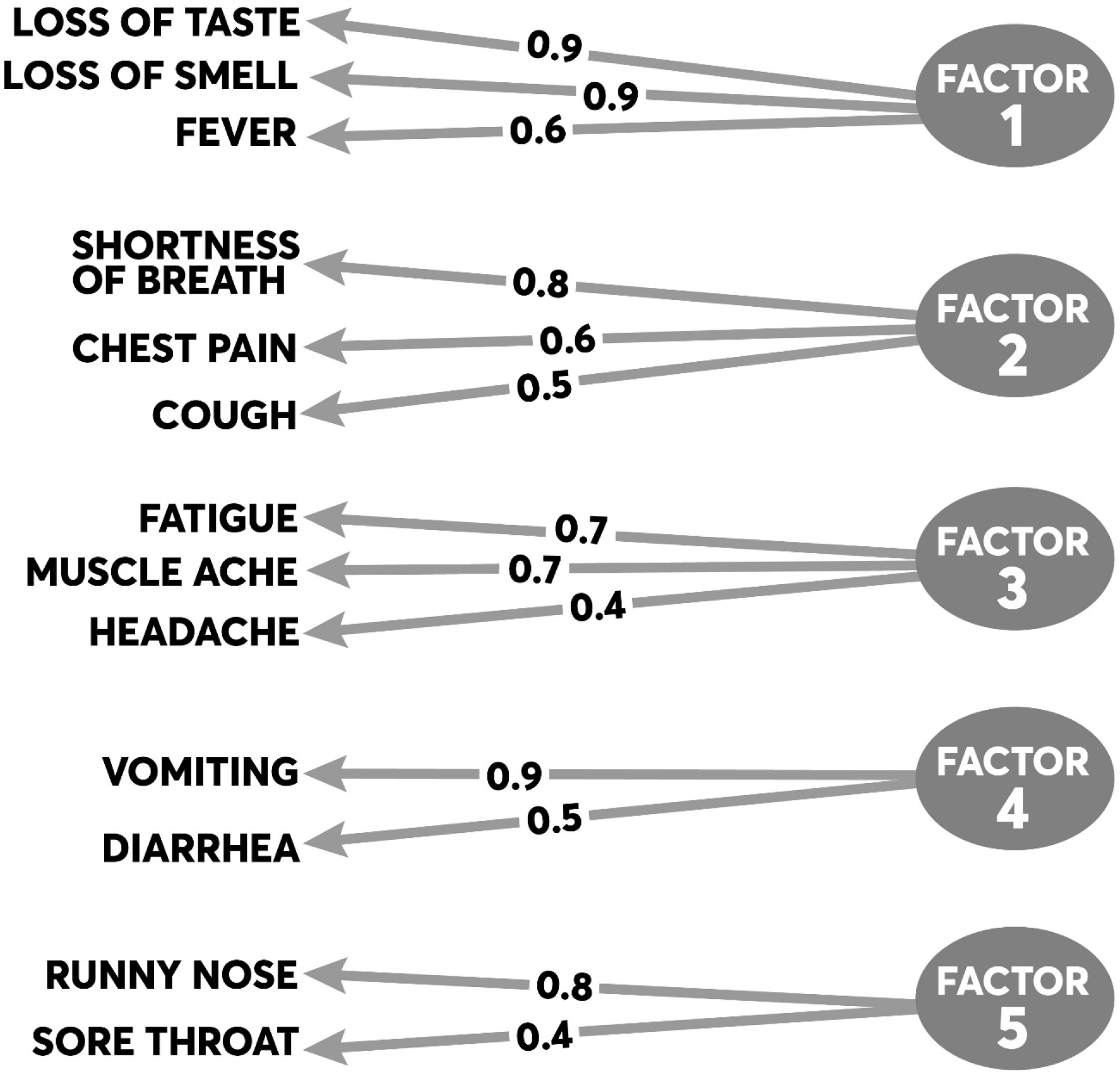
Results from the exploratory factor analysis of symptom clusters. **(caption)** The figure summarizes the principal symptoms loading on each factor. The proportions represent individual loadings for each symptom onto its factor based on the factor’s correlation matrix.

### Predicting COVID-19 Positivity based on Symptoms and Demographics

The results of the logistic regression model that examined how individual symptoms were associated with RT-PCR positivity are presented in **Table 3**. When controlling for demographics as well as testing group (e.g., random vs. nonrandom sample) and other symptoms, individual symptoms most strongly associated with high odds of RT-PCR SARS-CoV-2 positivity were fever (OR=5.34, p<0.001), anosmia (OR=4.08, p<0.001), ageusia (OR=2.38, p=0.006), and cough (OR=2.86, p<0.001). The ROC curve generated by a test based on this cluster of symptoms is presented in **Figure 2**. The AUC for the diagnostic index generated by the model which included this symptom cluster was 0.909. Individuals from the nonrandom sample were more likely than randomly selected participants to test positive (OR=9.34, p<0.001). Hispanic participants were more likely (OR=2.44, p<0.001) to test positive than non-Hispanic participants. Older participants, age ≥60 years, were less likely (OR=0.43, p<0.001) to test positive compared to participants under 40 years of age.

**Table 3.** Logistic regression model to predict SARS-CoV-2 RT-PCR positivity using participant reported symptoms and demographics.

| Symptom | AOR<sup>I</sup> | 95% CI<sup>I</sup> | p-value |
| --- | --- | --- | --- |
| **Fever** | 5.34 | 3.51, 8.12 | <0.001 |
| **Loss of Smell**<br>**(Anosmia)** | 4.08 | 2.14, 7.74 | <0.001 |
| **Loss of Taste (Ageusia)** | 2.38 | 1.28, 4.45 | 0.01 |
| **Cough** | 2.86 | 2.06, 3.97 | <0.001 |
| **Shortness of Breath** | 0.61 | 0.36, 1.00 | 0.05 |
| **Chest Pain** | 1.00 | 0.60, 1.64 | >0.999 |
| **Muscle Ache (Myalgia)** | 1.07 | 0.70, 1.61 | 0.75 |
| **Fatigue** | 1.25 | 0.84, 1.82 | 0.27 |
| **Headache** | 0.75 | 0.52, 1.07 | 0.12 |
| **Diarrhea** | 1.00 | 0.62, 1.58 | 0.99 |
| **Vomiting** | 1.35 | 0.47, 3.60 | 0.56 |
| **Sore Throat** | 0.80 | 0.53, 1.18 | 0.27 |
| **Runny Nose** | 1.14 | 0.79, 1.63 | 0.46 |
| **Demographics** |  |  |  |
| Age: <40 | — | — |  |
| Age: 40-59 | 0.88 | 0.66, 1.18 | 0.41 |
| Age: 60+ | 0.43 | 0.28, 0.63 | <0.001 |
| **Male** | 1.19 | 0.91, 1.56 | 0.21 |
| **Race** |  |  |  |
| White | — | — |  |
| Black or African<br>American | 0.80 | 0.49, 1.26 | 0.34 |
| Other | 1.68 | 1.17, 2.41 | 0.01 |
| **Hispanic** | 2.44 | 1.68, 3.54 | <0.001 |
| **Geography** |  |  |  |
| Urban | — | — |  |
| Rural | 1.53 | 0.97, 2.37 | 0.06 |
| Rural/Mixed | 1.30 | 0.79, 2.06 | 0.29 |
| **Participation** |  |  |  |
| Random Sample | — | — |  |
| Nonrandom Sample | 9.35 | 6.59, 13.4 | <0.001 |
| <sup>1</sup> AOR = Adjusted Odds Ratio, CI = Confidence Interval |  |  |  |

**Figure 2.**
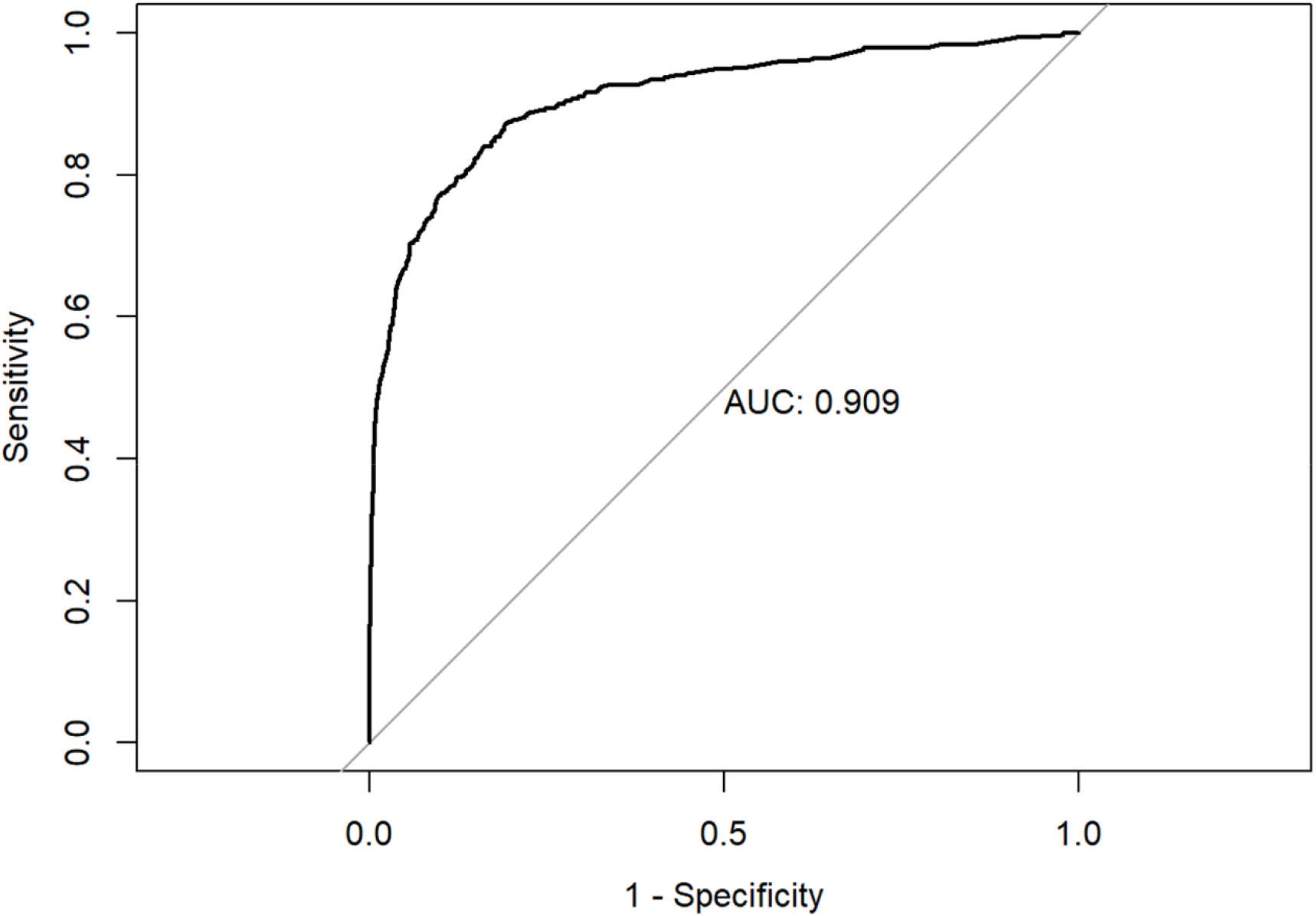
Area under the curve (AUC) for logistic model fit to predict RT-PCR status given symptoms or patient demographics.

## DISCUSSION

In this study of populations screened for COVID-19, we observed a wide constellation of symptoms present among those testing positive for active infection. Many symptoms commonly reported by infected participants (e.g., cough, fatigue, headache, myalgias) were similar to those identified by the CDC^1^ and those reported in prior studies.^3,7,8,11-13^ However, unlike prior studies, we also captured symptoms from those testing negative, enabling the examination of the sensitivity, specificity, PPV and NPV of individual symptoms and symptom combinations.

Our findings provide robust evidence to the growing body of studies that identify anosmia and ageusia as important symptoms of SARS-CoV-2 infection.^11,14,15^ These symptoms, especially in association with fever, are highly predictive of positive status and should trigger use of personal protective equipment (PPE), if universal PPE is not already being utilized, and prompt testing for COVID-19. Furthermore, particularly as influenza season arrives, it is important that health care providers specifically ask questions about loss of taste and smell in addition to traditional questions asked for assessing respiratory tract illness. Moreover, documenting these symptoms can aid public health officials in distinguishing COVID-19 from influenza within syndromic surveillance systems^16^ during flu season. Currently syndromes recommended by CDC and others overlap with influenza-like illness, and few electronic health record (EHR) systems capture information about loss of taste or smell.

We further observed a high rate of asymptomatic or pre-symptomatic individuals with 44.2% of the randomly selected participants and 20.2% of the nonrandom participants reporting a lack of related symptoms within two weeks of their positive RT-PCR test.^6^ A high rate of asymptomatic infection likely influenced the overall low PPV and sensitivity for individual symptoms and symptom combinations. The high proportion of asymptomatic cases complicates identification, control, and containment of new SARS-CoV-2 infections by public health authorities.

Virtual health care screening as well as chatbots and mobile applications^17,18^ are unlikely to refer asymptomatic individuals for testing. Without sufficient capacity for rapid, inexpensive testing, screening efforts may be hampered and accurate measurement of incidence and prevalence will be challenging.

As observed in other studies that analyzed cases,^19,20^ we found higher infection rates among Hispanic and non-white populations. These results were largely influenced by individuals involved in the nonrandom samples, who presented to community-based testing sites. Higher burden of disease among people of color have been widely reported and may stem from systemic inequities in economic, physical, and emotional health (e.g., social determinants).^21^ Targeted efforts to screen and test minority and other underserved populations, in coordination with public health and community-based organizations, are needed to adequately identify and reduce COVID-19 disparities.

### Limitations

We acknowledge several limitations. First, the two testing waves occurred as influenza season waned, which may decrease the presence of influenza-like-illness symptoms among those testing negative for COVID-19. Second, the association between age and RT-PCR positivity, e.g., older participants (60+ years) were less likely to test positive, may be driven by non-response to the sampling by these groups, as elderly and frail individuals may not have been as likely to participate in our studies as they were either tested in a nursing home (a population excluded from our sample) or were unlikely to present to a testing site. It is likely the differences observed in positivity among non-white and Hispanic populations are due to their self-selecting into testing at the nonrandom sites. They might have been more symptomatic and therefore motivated to seek testing as opposed to the randomized individuals. Finally, although the population was representative of Indiana, the cohort might not represent other states or the nation.

Despite these limitations, the study possessed significant strengths. The cohort was large and diverse, and it was drawn from a statewide population. Second, the symptoms examined were broad and included the most complete list based on available evidence. Finally, the robust methods yielded a strong model with face validity.

## CONCLUSION

This study finds that key symptoms for identifying active SARS-CoV-2 infection are anosmia and ageusia, especially in association with fever. Cough, especially when present along with shortness of breath and chest pain, is also an important symptom when diagnosing COVID-19. When laboratory testing is not accessible, these symptoms may help guide distinguishing COVID-19 from influenza-like illness. These symptoms should be used for screening and incorporated into clinical documentation to support public health identification of new cases and mitigation of disease spread.

## Data Availability

The data used in this study are available for review and potential use in secondary analyses from the IU Richard M. Fairbanks School of Public Health. Those interested in the data should contact the corresponding author for details as a data use agreement may be required prior to release.

## FUNDING

This work was supported by a grant from the State of Indiana to the IU Fairbanks School of Public Health to conduct seroprevalence testing in the state population. Dr. Dixon receives funding from the U.S. National Library of Medicine (T15LM012502) as well as the U.S. Centers for Disease Control and Prevention (U18DP006500) and the Indiana State Department of Health to support disease surveillance research.

## ACKNOWLEDGMENTS

The State of Indiana provided funding for the statewide COVID-19 surveillance testing study from which these data were derived. We thank our collaborators at the Indiana State Department of Health and the Governor’s Office as well as the Family Social Services Administration for their support of the statewide testing study and data management that enabled this research. The funder did not have influence over the findings or this publication. None of the authors have an association that might pose a conflict of interest.

## Notes

### Competing Interest Statement

The authors have declared no competing interest.

### Author Declarations

This study was determined to be exempt by the Institutional Review Board (IRB) at Indiana University under the public health surveillance exception.

### Summary of Updates

Updated the title and abstract to emphasize more the novelty of the paper and its methods. Further refined the text to emphasize the key findings. We seek to better distinguish this paper from others on COVID symptoms and signs.

